# Economic Gains from Health Equity: A Model for Pregnancy and Childbirth Policy Decisions

**DOI:** 10.1101/2024.10.21.24315689

**Authors:** Mark I. Evans, Gregory F. Ryan, Lawrence D. Devoe, George M. Mussalli, David W. Britt, Jaqueline M. Worth, Myriam Mondestin-Sorrentino, Christian R. Macedonia

**Affiliations:** Department of Obstetrics, Gynecology, and Reproductive Sciences, Icahn School of Medicine at Mount Sinai, NY; Fetal Medicine Foundation of America, NY; Department of Obstetrics & Gynecology, Yong Yoo Lin School of Medicine, National University of Singapore; Department of Obstetrics & Gynecology, Medical College of Georgia at Augusta University; Princeton Perinatal Institute, Lawrenceville, NJ; Lancaster Maternal Fetal Medicine, Lancaster PA

**Keywords:** maternal mortality ratio, neonatal mortality rate, disparities of care, prenatal care, maternal health deserts, health care economics, benefit-cost ratio, Value of a Statistical Life, Affordable Care Act, social determinants of health, economic integration, “DEVELOP”

## Abstract

**Importance:** American maternal and neonatal mortality rates are the worst of the world’s high-income countries. These rates are particularly low among patients of color, who have higher Cesarean delivery rates (CDR), higher healthcare costs, and poorer outcomes than White patients. However, common economic analyses do not address interlinked issues and therefore underestimate both the hidden causes of health inequities and the resultant costs to taxpayers. We have therefore designed a more comprehensive health economic model and metric (DEVELOP) that incorporates population health, equity, and economic integration.

**Design & Measures:** The DEVELOP model, a childbirth-specific model of the societal economic gain or loss related to healthcare outcomes, incorporates an individual’s long-term economic contributions into its calculations of economic benefits. We first used our model to estimate fiscal outcomes if each state’s CDR for Black patients was lowered to that of White patients. Second, we calculated the costs of “excess” CDR and mortalities among Black patients. Third, we incorporated the additional long-term economic contributions of mothers and their children.

**Results:** In the U.S., maternal and neonatal mortality rates and associated costs were higher for Black patients than White patients, and states with the lowest per capita health expenditures showed worse maternal outcomes and higher continuing costs. If the Black patient CDR were reduced to the White patient CDR, taxpayer-funded healthcare programs would save $263 million annually. Reducing the Black patient MMR would improve economic output by $224 million per year, and reducing the Black patient NMR would save $3.1 billion per year, for a combined economic improvement of $3.3 billion annually.

**Conclusions and Relevance:** The costs of improved prenatal care should be reconceptualized as investments for future economic growth rather than as short-term burdens. Policies blocking reasonable investments in health equity are counterproductive.

## INTRODUCTION

In the United States, racial disparities in health care outcomes pervade all aspects of medicine. In obstetrics, pregnancy care outcomes are commonly assessed using the maternal mortality ratio (MMR) and neonatal mortality rate (NMR).^1–15^ Here, we focus on integrative economic aspects of healthcare disparities in the United States, emphasizing the cost-effectiveness of hospital payments derived from taxpayer dollars. Over several decades, we have interacted with various federal and state governments concerning their medical/public health programs, and we have observed how legislators, agency directors, and staff typically evaluate funding requests.^16^ Unfortunately, governments often do not distinguish between medicine and other “special interest” groups seeking money from limited government resources.^16,17^

Taxpayer dollars already account for half of all healthcare spending in the U.S. Proposals for additional healthcare spending accordingly raise two important questions: 1) will the proposed program genuinely improve patient care; and 2) how much will implementation of these programs cost (or save)?^16,17^ Despite this need for clear cost-benefit analysis, typical econometric indices do not adequately measure the actual economic impacts or return on investment of such proposals. However, we cannot ignore the moral and scientific rationales by which government agencies should prioritize funding decisions.^18^

Numerous studies have detailed racial differences in obstetrical and neonatal outcomes.^1–15^ Unlike other medical problems, for which quantifying disparities requires multi-year timescales, most obstetric outcomes are quantifiable within a single year. One commonly used (albeit imperfect) measure of obstetrical care quality is the Cesarean delivery rate (CDR),^4,16^ which is uniformly higher for Black patients than for White patients. Cesarean deliveries carry an increased risk of morbidity and mortality during childbirth.^19^ Fortunately, the CDR is routinely calculated by numerous U.S. government agencies.^7,10–12^ We therefore used CDR benchmarks along with currently accepted U.S. government definitions for race to compare obstetric care between White and Black patient cohorts. We acknowledge that Hispanic and Asian patients represent sizable minorities in some states and will consider these groups in subsequent studies.

Minority women and children consistently have poorer outcomes.^1–16^ The higher CDR (and higher cost) among Black patients is partly due to the delayed initiation and reduced utilization and comprehensiveness of prenatal care.^7,10–12,19,20^ Strategies for promoting healthcare equity must confront the harsh economic realities of (i) the increased up-front costs required for enhanced care and (ii) the associated financial stresses on governmental and employer healthcare programs that must pay those costs.^16,17^

Achieving healthcare equity requires addressing a deeper set of problems at a systems level. To do so, we need better metrics and a common language to evaluate the impacts and costs of disparities in care. Here, we address this need by developing a new approach for quantifying the economic costs of healthcare disparities and then use this method to evaluate the economic consequences of higher CDRs, MMRs, and NMRs for Black patients in the United States.^7,10–12^ We additionally model, on a state-by-state basis, the economic consequences of hypothetically lowering the CDR for Black patients (through better care) to contemporaneous rates for White patients.

## MATERIALS AND METHODS

To quantify the additional costs for CD compared to vaginal delivery, we used data from multiple government-curated and well-vetted public repositories (Table 1). We calculated average additional costs associated with CDs vs. vaginal deliveries. We derived the “excess” number of CDs for Black patients per state by subtracting the total number of CDs performed for Black patients from the total number of CDs that would have been administered if the CDR for Black patients were equivalent to that of White patients.

**TABLE 1.**
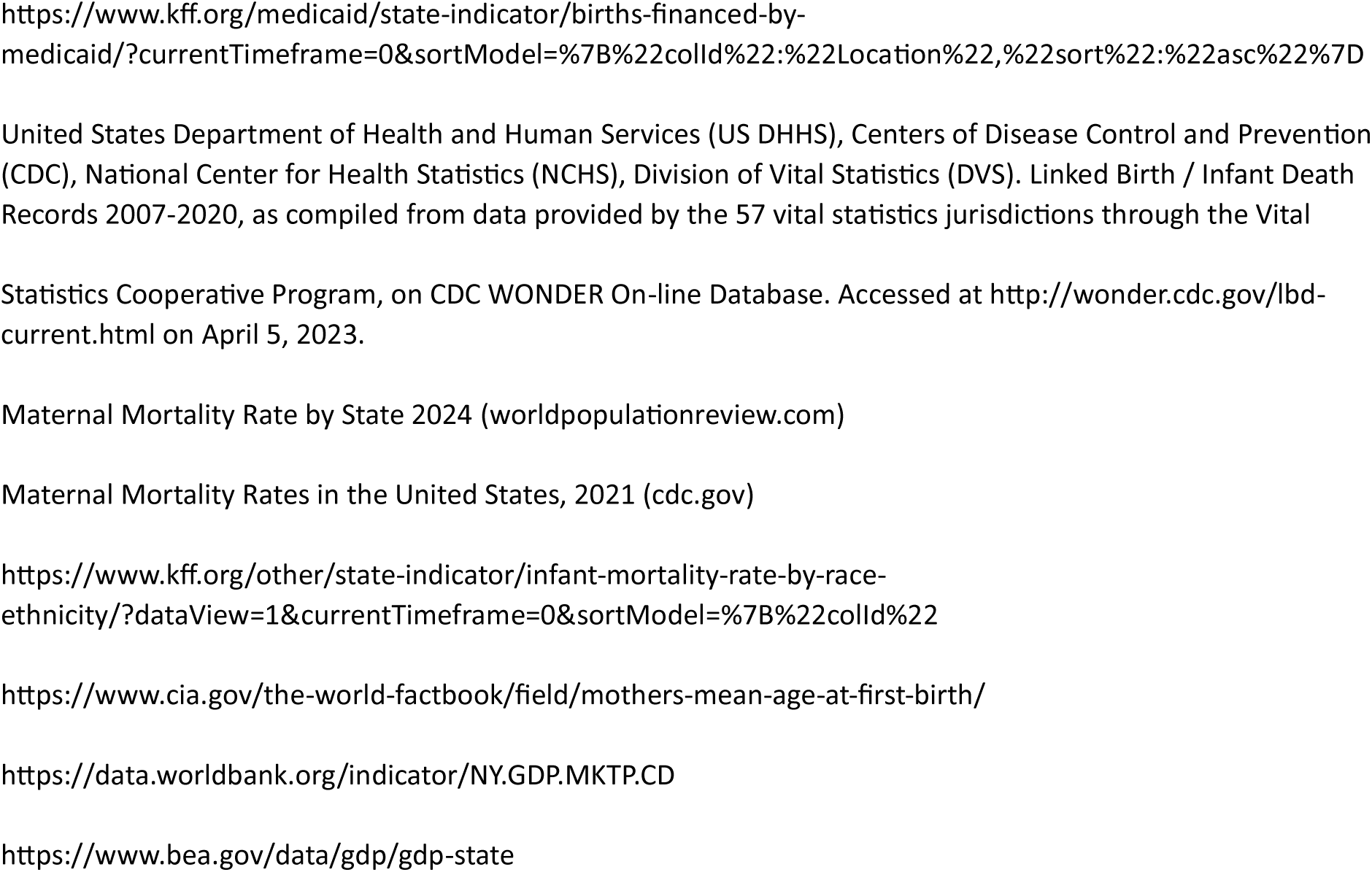
Databases Used.

We additionally calculated the financial consequences of different CDRs in each state using a gross domestic product (GDP)-based statistical dollar value of each life analogous to the “Value of a Statistical Life” developed by the National Bureau of Economic Research.^21–23^ The excess costs associated with a higher CDR were calculated as:

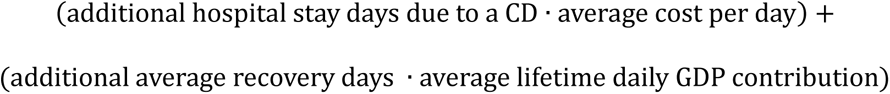

To quantify the economic benefits of reduced MMR and NMR, we propose a new economic model (**De**velopment of **V**alue and **E**conomics of **L**ife **O**ptimization and **P**reservation, [“DEVELOP”]), from which we calculate a “DEVELOP Score” using generally accepted accounting principles. The DEVELOP Score uses the current GDP per capita in the state (or country) of interest, the associated 10-year average annual GDP growth rate, and the average life expectancy (Figure 1). We chose GDP for our calculations because the UN, U.S., and many other nations and organizations use GDP as a standard measure of wealth and productivity. To calculate DEVELOP Scores in regions with a negative 10-year average annual GDP growth rate, we set the annual growth rate to 0%. Average life expectancy data come from the U.S. Census Bureau, World Bank, or UN. To determine the economic costs of maternal mortality in each state, we subtracted the average female life expectancy from the average age at childbirth to estimate years of lost life. This value was then multiplied by the annual GDP per capita. For neonatal mortality, we performed a similar calculation using the overall average life expectancy in each state.

**Figure 1:** Mathematical calculations used to determine the “DEVELOP” Score. Legend: Steps in the calculation of the DEVELOP Score using California data as an example.

We applied a present value discount rate of 10% to the GDP, though this can be adjusted up or down as specific circumstances might warrant. Higher discount rates lead to lower DEVELOP Scores, and vice versa. Purchasing power parity could be substituted for GDP for comparisons among countries.

We also added a disability-adjusted weight to the DEVELOP score to account for the economic consequences of different morbidities depending on their severity. For example, severe cerebral palsy (e.g., spastic quadriplegia) is weighted more heavily than moderate cases.^20,1^ CDs have excess costs from additional hospital and recovery days compared to uncomplicated vaginal deliveries.

Using this framework, we calculated DEVELOP scores nationwide and for every U.S. state for each year of lost life or year of disabled life averted (Table 2). For example, the 2020 World Bank statistics show that Mexico’s average per capita GDP was $10,045.68. The DEVELOP Score of newborns and mothers in Mexico correspondingly calculate to $114,993.69 and $113,728.88. As a second example, the U.S. per capita GDP is $70,248.63, and the corresponding DEVELOP Scores are $907,912.09 per newborn and $894,544.63 per mother. However, DEVELOP Scores vary widely among states: California has newborn and maternal DEVELOP Scores of $1,466,462.55 and $1,423,480.82, respectively, whereas neighboring Arizona is 40% lower at $1,001,930.44 and $962,099.64, respectively. Mississippi newborn and maternal DEVELOP Scores are even lower: $546,819.37 and $539,656.44, respectively.

**TABLE 2:** DEVELOP SCORE ANALYSIS. Legend: States ranked from best to worst based on their MMR, NMR, and combined score.

| State Name | 10 Year Average Per Capita Healthcare Spend | 10 Year Per Capita Healthcare Spend Rank | MMR Rank (lowest MMR = 1) | NMR Rank (lowest NMR = 1) | Statistical Value of a Mother's Life Based on GDP CAGR 2014-2022 | Statistical Value of a Neonate's Life Based on GDP CAGR 2014-2022 | Healthcare Rank (Lowest = 1) | Economic Rank (Lowest = 1) | Combined Rank (Lowest Score = 1) |
| --- | --- | --- | --- | --- | --- | --- | --- | --- | --- |
| Alabama | \$ 7,769.90 | 41 | 48 | 46 | \$ 689,549.67 | \$ 703,584.18 | 48 | 51 | 51 |
| Alaska | \$ 11,498.00 | 2 | 34 | 19 | \$ 856,269.57 | \$ 859,859.50 | 16 | 18 | 16 |
| Arizona | \$ 7,146.20 | 47 | 43 | 23 | \$ 962,099.64 | \$ 1,001,930.44 | 29 | 30 | 31 |
| Arkansas | \$ 7,826.10 | 40 | 50 | 49 | \$ 693,395.36 | \$ 705,813.03 | 49 | 45 | 48 |
| California | \$ 8,240.50 | 32 | 6 | 5 | \$ 1,423,480.82 | \$ 1,466,462.55 | 6 | 6 | 2 |
| Colorado | \$ 7,220.60 | 46 | 8 | 17 | \$ 1,295,198.07 | \$ 1,343,188.91 | 5 | 17 | 7 |
| Connecticut | \$ 10,509.60 | 7 | 12 | 12 | \$ 1,050,239.15 | \$ 1,067,239.22 | 25 | 14 | 19 |
| Delaware | \$ 10,865.30 | 6 | 18 | 13 | \$ 950,648.78 | \$ 959,721.25 | 18 | 15 | 15 |
| District of Colu | \$ 12,680.50 | 1 | 1 | 2 | \$ 3,099,718.65 | \$ 3,127,780.30 | 3 | 7 | 1 |
| Florida | \$ 8,471.80 | 28 | 36 | 30 | \$ 998,340.83 | \$ 1,038,533.53 | 42 | 16 | 29 |
| Georgia | \$ 7,138.60 | 48 | 49 | 38 | \$ 1,030,527.20 | \$ 1,068,057.68 | 47 | 24 | 40 |
| Hawaii | \$ 8,036.40 | 35 | 19 | 21 | \$ 769,658.49 | \$ 774,611.70 | 13 | 46 | 32 |
| Idaho | \$ 6,971.60 | 50 | 16 | 20 | \$ 937,344.22 | \$ 988,259.39 | 7 | 27 | 17 |
| Illinois | \$ 8,510.00 | 25 | 17 | 27 | \$ 979,688.59 | \$ 994,461.27 | 24 | 32 | 27 |
| Indiana | \$ 8,626.70 | 22 | 41 | 45 | \$ 829,514.84 | \$ 847,542.55 | 40 | 28 | 38 |
| Iowa | \$ 8,360.30 | 31 | 21 | 15 | \$ 841,931.40 | \$ 859,236.74 | 15 | 44 | 33 |
| Kansas | \$ 7,913.70 | 38 | 27 | 41 | \$ 914,318.15 | \$ 936,637.96 | 32 | 38 | 39 |
| Kentucky | \$ 8,501.90 | 26 | 45 | 39 | \$ 714,911.16 | \$ 728,646.26 | 46 | 40 | 45 |
| Louisiana | \$ 8,482.80 | 27 | 46 | 50 | \$ 609,150.49 | \$ 612,797.47 | 50 | 48 | 49 |
| Maine | \$ 9,978.80 | 10 | 3 | 36 | \$ 841,553.90 | \$ 867,347.52 | 20 | 10 | 12 |
| Maryland | \$ 9,176.40 | 17 | 23 | 34 | \$ 919,758.58 | \$ 934,313.47 | 36 | 31 | 37 |
| Massachusetts | \$ 11,266.00 | 3 | 9 | 4 | \$ 1,376,581.06 | \$ 1,413,648.63 | 10 | 2 | 3 |
| Michigan | \$ 8,517.40 | 24 | 20 | 44 | \$ 781,338.10 | \$ 801,458.94 | 37 | 42 | 43 |
| Minnesota | \$ 9,372.20 | 16 | 5 | 11 | \$ 972,615.17 | \$ 994,432.19 | 4 | 12 | 6 |
| Mississippi | \$ 7,954.30 | 36 | 51 | 51 | \$ 539,656.44 | \$ 546,819.37 | 51 | 47 | 50 |
| Missouri | \$ 8,430.00 | 29 | 32 | 33 | \$ 763,128.49 | \$ 776,435.81 | 31 | 43 | 41 |
| Montana | \$ 8,568.40 | 23 | 38 | 16 | \$ 739,865.20 | \$ 756,421.46 | 22 | 33 | 23 |
| Nebraska | \$ 8,814.10 | 20 | 33 | 29 | \$ 1,057,313.59 | \$ 1,087,031.21 | 26 | 5 | 13 |
| Nevada | \$ 7,137.10 | 49 | 26 | 18 | \$ 1,024,280.51 | \$ 1,056,867.29 | 30 | 25 | 24 |
| New Hampshire | \$ 10,080.00 | 9 | 7 | 3 | \$ 1,016,798.18 | \$ 1,051,117.95 | 11 | 3 | 5 |
| New Jersey | \$ 9,744.80 | 14 | 30 | 6 | \$ 1,004,362.80 | \$ 1,022,950.46 | 23 | 9 | 14 |
| New Mexico | \$ 7,552.30 | 44 | 42 | 24 | \$ 679,609.81 | \$ 688,150.84 | 27 | 50 | 42 |
| New York | \$ 10,907.90 | 5 | 25 | 10 | \$ 1,375,330.36 | \$ 1,399,409.19 | 28 | 1 | 11 |
| North Carolina | \$ 7,718.60 | 42 | 24 | 43 | \$ 957,599.03 | \$ 985,636.94 | 33 | 22 | 25 |
| North Dakota | \$ 9,572.10 | 15 | 29 | 22 | \$ 927,360.27 | \$ 936,010.64 | 17 | 26 | 20 |
| Ohio | \$ 8,926.70 | 19 | 28 | 42 | \$ 868,808.55 | \$ 887,554.70 | 35 | 29 | 36 |
| Oklahoma | \$ 8,053.10 | 33 | 40 | 35 | \$ 659,769.86 | \$ 666,520.24 | 41 | 49 | 47 |
| Oregon | \$ 8,426.10 | 30 | 13 | 9 | \$ 1,106,935.23 | \$ 1,149,145.91 | 8 | 4 | 4 |
| Pennsylvania | \$ 9,846.40 | 13 | 15 | 25 | \$ 855,378.75 | \$ 870,131.80 | 21 | 34 | 26 |
| Rhode Island | \$ 9,953.30 | 12 | 14 | 8 | \$ 763,108.89 | \$ 773,673.09 | 19 | 37 | 28 |
| South Carolina | \$ 7,560.70 | 43 | 44 | 40 | \$ 792,797.70 | \$ 816,523.26 | 45 | 41 | 46 |
| South Dakota | \$ 9,970.40 | 11 | 37 | 47 | \$ 917,945.05 | \$ 939,701.86 | 34 | 11 | 21 |
| Tennessee | \$ 7,937.90 | 37 | 47 | 37 | \$ 994,681.51 | \$ 1,028,857.27 | 43 | 20 | 35 |
| Texas | \$ 7,239.60 | 45 | 35 | 26 | \$ 1,128,559.30 | \$ 1,159,073.98 | 39 | 19 | 30 |
| Utah | \$ 6,299.40 | 51 | 11 | 28 | \$ 1,288,664.82 | \$ 1,398,271.25 | 9 | 13 | 8 |
| Vermont | \$ 11,021.30 | 4 | 2 | 1 | \$ 754,792.99 | \$ 769,363.79 | 1 | 21 | 9 |
| Virginia | \$ 7,883.50 | 39 | 39 | 32 | \$ 969,834.98 | \$ 990,950.21 | 38 | 23 | 34 |
| Washington | \$ 8,040.90 | 34 | 22 | 14 | \$ 1,616,822.41 | \$ 1,696,692.74 | 14 | 8 | 10 |
| West Virginia | \$ 10,270.70 | 8 | 31 | 48 | \$ 596,718.93 | \$ 603,629.62 | 44 | 39 | 44 |
| Wisconsin | \$ 8,744.60 | 21 | 4 | 31 | \$ 828,139.55 | \$ 848,393.47 | 12 | 36 | 22 |
| Wyoming | \$ 8,935.60 | 18 | 10 | 7 | \$ 806,332.13 | \$ 811,695.11 | 2 | 35 | 18 |
| United States | | | | | \$ 957,648.34 | \$ 976,725.33 | | | |

## RESULTS

In all 50 states and Washington D.C., the CDR was higher for Black patients than for White patients (Figure 2). Nationally, the Black patient CDR was 36.8%, compared to 31.0% for White patients. For comparison, the CDR in sub-Saharan Africa is 7.1%.^24^ If the 2021 CDR for Black patients was equivalent to that of White patients, insurance companies and taxpayer-funded health programs would have saved $263 million. Likewise, reducing the Black patient MMR from 69.9 to the White patient MMR of 26.6 would have increased economic output by $224 million, and reducing the Black patient NMR from 10.38 to the White patient NMR of 4.40 would have added $3.1 billion back to the economy.

**Figure 2.** Racial variation in CDR, MMR, and NMR, and their financial implications. Legend: Black patients have increased rates of Cesarean deliveries, maternal mortality, and neonatal mortality relative to White patients in all U.S. states. Our model suggests that lowering the Black patient CDR and MMR/NMR to that of White patients through the provision of better care would save $213 million and $3 billion per cohort-year, respectively, for a total of approximately $3.5 billion in economic improvement.

By our definitions, racial disparities in these three healthcare outcomes cost the U.S. a total of $3.3 billion annually (Figure 2). If, by better provision of obstetric and neonatal care, the CDR, MMR, and NMR of Black patients equaled those of White patients, the U.S. economy would improve by $3.522 billion per cohort-year. This savings would be $471 per delivery in California and $1,665 in Georgia (Figure 2).

## DISCUSSION

Of all the world’s high-income countries, the U.S. has the worst maternal and neonatal outcome statistics, and healthcare costs and outcomes differ substantially among racial groups.^25^ This paradox of a high GDP but low-quality obstetric care requires deeper analysis. Indeed, if these racial disparities in care were equalized, the savings could be reallocated to other approaches and programs for improving overall healthcare.^26^ Resolving such discrepancies is a continual, complex process addressed extensively elsewhere.^27–32^

Restrictions that limit patient access to care can be either physical (e.g., hospital closures, staff reductions) or programmatic (e.g., indicated vs. actual access). Explicit bias (e.g., overt racism) and implicit bias (e.g., physician acquiescence to corporate imperatives for financial surplus) represent additional ‘mental’ or ‘moral’ restrictions which, while important, are methodologically more difficult to quantify.

We have intentionally focused on developing economic measures that encompass a longer time frame. The costs of improving care can eventually be recovered; in addition to the huge non-financial gain of every human life preserved, patients with improved health outcomes will make larger continuing contributions to the GDP.

One of the most significant barriers to developing a unified strategy for improving obstetric (and other) care is that the various healthcare stakeholders have different economic priorities:

1. **Hospitals** focus on the immediate costs of care. Compensation from insurance companies and governmental agencies affects their bottom line and future viability.
2. **States** administer Medicaid programs and focus on the costs and revenues of healthcare programs as part of their annual budgets. However, the immediate costs of providing obstetric care should be compared to resultant costs of providing neonatal and infant care. Because many states require statutory balanced budgets, the financial consequences of disparities in care can vary considerably from year to year.
3. **The federal government,** through its budget process and the Federal Reserve, supplies money to the economy. This responsibility allows the federal government to consider a longer, multi-year timeframe. Nevertheless, federal programs generally do not emphasize the long-term financial consequences of healthcare disparities and their attendant excess costs.^16,17^ The federal bureaucracy has traditionally prioritized funding for seniors over women and children, reflecting the historical strength of this voting bloc.

Individual projections are always fraught with uncertainties, but cost-benefit estimates generated on a population basis could be more precisely refined. Our data suggest that reframing economic timelines and horizons in healthcare cost-benefit analysis could justify more up-front investments to reduce the costs incurred at birth and beyond. Such investments would improve both public and private bottom lines not only by reducing maternal and neonatal costs but also by providing long-term financial and social contributions that are not captured in a hospital’s profit and loss statements. Indeed, our estimate of an annual cost savings of $3.4 billion is likely a significant underestimation of the true return on investment.

Healthcare disparities cannot be cured by governmental fiats; improvements must instead be evaluated as part of a multi-step, contextualized process. By creating sub-goals and evaluating them “tranche by tranche,” like responsible businesses do, stakeholders and policymakers could generate greater enthusiasm for better care and improved outcomes.

The challenge in this process of improvement has been finding suitable, quantitative metrics that compare service costs and their resulting economic benefits. We believe the commonly used “benefit cost ratio” and the “social rate of return” developed at the University of Chicago^22^ are suboptimal for healthcare economics because they focus primarily on social rather than financial outcomes. The U.S. Environmental Protection Agency and the U.S. Department of Transportation use Vanderbilt University Law School’s “Value of a Statistical Life,”^23^ which, despite its utility, ignores too many important economic variables to be reasonably precise. Neumann’s cost effectiveness thresholds use quality-adjusted life years, which many experts believe have little theoretical or empirical foundation.^33^

To determine the cost efficiency of an action, economists typically employ evaluative tools such as cost-effective analysis or the incremental cost effectiveness ratio. The equivalent tools for quantifying treatment outcomes include disability-adjusted life years, quality-adjusted life years, averted years of life lost, and averted years of life lost due to disability. However, these metrics do not adequately address direct economic questions because they mostly compare the costs of the proposed services to the cost of the “next best alternative.” We believe that the overall focus of healthcare cost-benefit analysis should be on increasing long-term economic value, exemplified by the question “how much does a healthy individual add to the economic output of a state or nation?” This question is different from the work requirements to ensure a motivated workforce. Research has shown that healthy individuals and communities are more economically productive in multiple ways.^34^ We cannot ignore the fact that differing approaches to the costs of care must be justified in a market economy.

We therefore created DEVELOP, which is a simplified version of the Value of a Statistical Life. We acknowledge that putting dollar values on human life is morally distasteful, especially when done to avoid ethical responsibilities. Our approach and acronym are therefore directed towards “life optimization and preservation,” framing the analysis on integrity and noble principles. Importantly, our valuations yield concrete numbers—dollars of economic benefits associated with every life saved or morbidity prevented and the specific cost benefits for stakeholders.^16,17^

In the DEVELOP model, the societal economic benefits of preventing morbidities and mortalities include the increased tax revenue generated from an individual’s lifetime earnings. The longer someone lives without disability, the more economically productive that individual becomes, and the more tax revenue they generate.^35^ Later versions of our model will consider secondary economic benefits realized through an individual’s purchases of goods (food, housing, etc.) or services (education, etc.). Furthermore, a reduced MMR means that the state, family, and/or community allocates less time and resources to helping raise a deceased woman’s children, thereby creating more economic opportunities. Reducing the CDR similarly lowers costs and frees hospital beds. Every dollar saved with better labor and delivery management increases the capacity of the health care system to provide improved healthcare.

Modeling the effect of CDR reduction for Black patients, as we did here, is only a starting point for the DEVELOP approach. Indeed, DEVELOP provides a quantitative, contemporaneous analytic metric. However, we are not suggesting that the current CDR for White patients is a “good” number, especially in comparison to other G7 nations.^36^ We are instead developing methods designed to safely guide policy on lowering the CDR, which, by extension, should have disproportionate benefits for patients of color and rural patients of all ethnicities.^28,37,38^

In the U.S., most childbirth deliveries are financed by either Medicaid or indemnity insurance companies; the latter negotiates fees. Administrators for the Centers for Medicare and Medicaid Services (CMS) are the fiduciaries for physician reimbursement, supposedly tailoring the Medicare/Medicaid fee tables to address historic underpayments for women’s medicine relative to more male-oriented specialties like urology, cardiology, and orthopedics.^39,40^ The CMS payment tables come from an American Medical Association working group called the Relative Value Scale Update Committee (RUC). Although women and children comprise 62% of the U.S. population, Pediatrics and Obstetrics/Gynecology only have two of the 32 specialty seats on this committee. It is therefore unsurprising that pediatrics and women’s health are perennially underfunded.

The emotional benefits of better care are real and important, but they cannot be calculated with current financial metrics. Indeed, typical financial metrics have not moved the needle in any substantial way: U.S. healthcare budgets still skew strongly toward conditions affecting older men.^39,40^ Our novel approach attempts to correct some of these gaps.

Better care means that hospitals can more efficiently use and manage their finite bed capacities, thereby increasing healthcare throughput. Moreover, healthcare agencies can reduce expenditures on the otherwise lifelong costs of care for disabled children. Social Security Disability Insurance monthly payments, over and above the medical costs of care, average $1,537.00/month from 18 years of age until death (January 2024 data).^41^

Educating a disabled child through out-of-district placement (ages of 4-22 for up to 18) costs up to $500,000 per year.^42^ Liability insurance companies have exposures from bad outcomes including the costs of physical therapy, medications, rehabilitation, doctors’ visits, surgeries, testing, and procedures. From the taxpayer perspective, a more efficient system could potentially reduce the tax burden of care for disabled children and individuals.

From the public policy and technology assessment perspectives, proposals for increasing up-front healthcare expenditures encounter considerable resistance, especially from states that have traditionally minimized their safety net programs. In these environments, the process of reconceptualizing expenses needs to be evidence-based: for example, the high initial costs involved in starting a business or buying a home are often justified because the value of these entities should be much higher decades into the future. To shift the prevailing narrative of healthcare costs, we must reframe commonly accepted paradigms. Only then can public policies be effectively implemented.^43^

Some inertia in the system is always present but difficult to quantify.^44^ Public distrust in a system that, for hundreds of years, created barriers to and lower tiers of care cannot instantly vanish. Trust must be earned through repeated acts of trustworthiness. This lack of trust partially explains why so many patients at the highest end of the need spectrum do not take advantage of supposedly available healthcare services. Inequities in health access and availability clearly remain.

Our data, along with other studies, show that methods for implementing and diffusing healthcare services must be improved to address disparities of care and make sure healthcare services are equitably available.^345,46^ Compartmentalizing medical expenditures would permit a continuing analysis of the overall economic value of healthcare, which accounts for 17% of the U.S. GDP. Indeed, the federal government pays more than 50% of U.S. healthcare costs and has a predominant role in setting healthcare prices. However, in our opinion, CMS has not provided sufficient incentives for health insurers, hospitals, and independent medical practices to improve maternal outcomes.

Policy and budget initiatives have improved some components of healthcare but have not yet meaningfully overcome the large, systemic biases that contribute to current inequities in adverse healthcare outcomes. Only when these issues receive specific attention from diverse stakeholders will we see real progress for the large patient groups that have historically been adversely affected.

### Strengths and limitations

We believe our work is the first study to comprehensively model both short- and long-term economic consequences of how governments and private insurers allocate resources for obstetrical and resultant neonatal/infant care. Although our conclusions may seem intuitively obvious, a deep search in PubMed suggests that ours is the first research paper to support these intuitive beliefs with factual data and analytics. We hope that our data-driven analysis of the economic consequences of healthcare disparities will engender public policy debates that focus on timelines longer than the next election cycle. We expect future versions of the DEVELOP model to offer even deeper insights.

Despite this major step forward, our results do not identify any universal theme underlying the wide variations in (i) health care expenditures among state indemnity and Medicaid programs, (ii) approvals for services by managed care gatekeepers, and (iii) what different companies and governments consider adequate treatments. Indeed, new approaches to care continually transition from “experimental” to routine, eventually (and often too slowly) making them eligible for future insurance coverage.^44,47^

Deeper analyses will also be required to determine how governments create “winners,” whereby large health systems have large profit margins for services such as neonatal intensive care even as the overall health universe loses.^48,50^ Even so, resistance to changing approaches to care only hinders the health care landscape from taking advantage of new methods to amplify the voices of historically underserved demographic groups.

### Conclusions

Miserliness in the provision of obstetrical care does not save money in either the short or long term: instead, lower “up front” expenditures ultimately cost the U.S. more than $3 billion per year in direct outlays and tens of billions of dollars amortized due to lost years of life and productivity. We believe that the challenge of improving disparities in maternal, fetal, and infant care should be reconceptualized to include a larger, overarching viewpoint that considers both medicine and economics in long-term timescales.^40^ Better economic evaluations will empower concomitant improvements in both data evaluation and machine learning approaches to support more sophisticated analyses and program development. ^51–54^ Under the framework we propose here, the justifications for enhancing the provision of prenatal care become economically, ethically, morally, and practically compelling.

## DECLARATIONS

### DATA SHARING STATEMENT

All data were taken from publicly available databases and then analyzed: we have aggregate, de-identified statistics but no individual patient data. When our current studies based on this data have been completed (no specific date yet established), the aggregate data will be publicly shared in the form of data tables on the lead author’s website.

### FUNDING

We received no outside funding for this project or manuscript.

### IRB APPROVAL

Because this project has no individual patient data and is only a secondary analysis of previously published, de-identified data, no IRB approval is necessary.

### RACE and ETHNICITY

Data on racial differences are taken directly from contemporaneous U.S. government databases using their definitions. We have not attempted to dissect or alter these definitions.

DECLARATION of AI and AI-assisted technologies in the writing process:

No AI or assisted technologies were used in the preparation of this manuscript.

## Data Availability

All data produced in the present study are available upon reasonable request to the authors

https://www.kff.org/medicaid/state-indicator/births-financed-by-medicaid/?currentTimeframe=0&sortModel=%7B%22colId%22:%22Location%22,%22sort%22:%22asc%22%7D

https://www.cdc.gov/nchs/nvss/linked-birth.htm

http://wonder.cdc.gov/lbd-current.html

https://worldpopulationreview.com/

https://www.cdc.gov/nchs/data/hestat/maternal-mortality/2021/maternal-mortality-rates-2021.htm

https://www.kff.org/other/state-indicator/infant-mortality-rate-by-race-ethnicity/?dataView=1&currentTimeframe=0&sortModel=%7B%22colId%22

https://www.cia.gov/the-world-factbook/field/mothers-mean-age-at-first-birth/

https://data.worldbank.org/indicator/NY.GDP.MKTP.CD

https://www.bea.gov/data/gdp/gdp-state

## Notes

### Competing Interest Statement

The authors have declared no competing interest.

### Funding Statement

This study did not receive any funding

### Author Declarations

The study used (or will use) ONLY openly available human data that were originally located at: https://www.kff.org/medicaid/state-indicator/births-financed-by-medicaid/?currentTimeframe=0&sortModel=%7B%22colId%22:%22Location%22,%22sort%22:%22asc%22%7D United States Department of Health and Human Services (US DHHS), Centers of Disease Control and Prevention (CDC), National Center for Health Statistics (NCHS), Division of Vital Statistics (DVS). Linked Birth / Infant Death Records 2007-2020, as compiled from data provided by the 57 vital statistics jurisdictions through the Vital Statistics Cooperative Program, on CDC WONDER On-line Database. Accessed at http://wonder.cdc.gov/lbd-current.html on April 5, 2023. Maternal Mortality Rate by State 2024 (worldpopulationreview.com) Maternal Mortality Rates in the United States, 2021 (cdc.gov) https://www.kff.org/other/state-indicator/infant-mortality-rate-by-race-ethnicity/?dataView=1&currentTimeframe=0&sortModel=%7B%22colId%22 https://www.cia.gov/the-world-factbook/field/mothers-mean-age-at-first-birth/ https://data.worldbank.org/indicator/NY.GDP.MKTP.CD https://www.bea.gov/data/gdp/gdp-state

## REFERENCES

1. Declercq E, Barnard-Mayers R, Zephyrin LC, Johnson K. The U.S. maternal health divide: the limited maternal health services and worse outcomes of states proposing new abortion restrictions. The Commonwealth Fund. 2024. https://www.commonwealthfund.org/publications/issue-briefs/2022/dec/us-maternal-health-divide-limited-services-worse-outcomes

2. Declercq E, Thoma M. Measuring US maternal mortality. JAMA. 2023;330(18):1731–1732. doi:10.1001/jama.2023.19945

3. Gunja MZ, Gumas ED, Williams RDI. The U.S. maternal mortality crisis continues to worsen: an international comparison. The Commonwealth Fund. 2024. https://www.commonwealthfund.org/blog/2022/us-maternal-mortality-crisis-continues-worsen-international-comparison

4. Leonard SA, Berrahou I, Zhang A, Monseur B, Main EK, Obedin-Maliver J. Sexual and/or gender minority disparities in obstetrical and birth outcomes. Am J Obstet Gynecol. 2022;226(6):846.e1–846.e14. doi:10.1016/j.ajog.2022.02.041

5. Osterman MJK, Hamilton BE, Martin JA, Driscoll AK, Valenzuela CP. Births: Final Data for 2021. Natl Vital Stat Rep. Jan 2023;72(1):1–53.

6. Corbisiero MF, Tolbert B, Sanches M, et al. Medicaid coverage and access to obstetrics and gynecology subspecialists: findings from a national mystery caller study in the United States. Am J Obstet Gynecol. 2023;228(6):722.e1–722.e9. doi:10.1016/j.ajog.2023.03.004

7. Ely DM, Driscoll AK. Infant Mortality in the United States, 2020: Data From the Period Linked Birth/Infant Death File. Natl Vital Stat Rep. Sep 2022;71(5):1–18.

8. Crockett AH, Chen L, Heberlein EC, et al. Group vs traditional prenatal care for improving racial equity in preterm birth and low birthweight: the Centering and Racial Disparities randomized clinical trial study. Am J Obstet Gynecol. 2022;227(6):893.e1–893.e15. doi:10.1016/j.ajog.2022.06.066

9. Kern-Goldberger AR, Malhotra T, Zera CA. Society for Maternal-Fetal Medicine Special Statement: Utilizing telemedicine to address disparities in maternal-fetal medicine: a call to policy action. Am J Obstet Gynecol. 2024;230(5):B6–B11. doi:10.1016/j.ajog.2023.11.001

10. Davidson KW, Terry MB, Braveman P, Reis PJ, Timmermans S, Epling JW, Jr. Maternal mortality: A National Institutes of Health Pathways to Prevention panel report. Obstet Gynecol. 2024;143(3):e78–e85. doi:10.1097/AOG.0000000000005488

11. Arechvo A, Nikolaidi DA, Gil MM, et al. Maternal race and stillbirth: cohort study and systematic review with meta-analysis. J Clin Med. 2022;11(12). doi:10.3390/jcm11123452

12. McIlwraith C, Sanusi A, McGwin G, Jr., Battarbee A, Subramaniam A. Recurrent severe maternal morbidity in an obstetric population with a high comorbidity burden. Obstet Gynecol. 2024;143(2)

13. Goldfarb S, Deichen Hansen M, Day J, Brown Speights JS, Rust G, Harman J: State variation in progress towards eliminating disparities in Infant Mortality, 2007-19. Health Affairs 2024;43

14. Brown Speights JS, Goldfarb SS, Wells BA, Beitsch L, Levine RS, Rust G. State-level progress in reducing the Black-White infant mortality gap, United States, 1999–2013. Am J Public Health. 2017;107(5):775–82.

15. Wallace M, Crear-Perry J, Richardson L, Tarver M, Theall K. Separate and unequal: structural racism and infant mortality in the US. Health Place. 2017;45:140–4.

16. Evans MI, Hanft R. The introduction of new technologies. ACOG; 1997:3.

17. Cohen A, Hanft R, Encinosa W, Spernak S, Stewart S, White C. Technology in American health care: policy directions for effective evaluation and management University of Michigan Press; 2004.

18. Buettgens M, Blavin F, Pan C: The affordable care act reduced income inequality in the US. Health Aff. 2021 40:121–129

19. Evans MI, Ryan GF, Mussalli G, Britt DW, Worth J, Devoe LD: Another Reason to Address Social Determinants and Disparities: It Saves Money. Am J Obstet Gynecol 2024;230:S100

20. Evans M, Ryan G, Britt D, et al. Reframing the economic arguments for improving obstetrical care: saving money costs too much. presented at: 71st Society for Reproductive Investigation Scientific Meeting; March 12-16 2024; Vancouver, BC, Canada.

21. Artiga S, Hinton E. Beyond health care: the role of social determinants in promoting health and health equity 2018. https://files.kff.org/attachment/issue-brief-beyond-health-care

22. Kremer M, Gallant S, Rostapshova O, et al. Is development innovation a good investment? Which innovations scale? Evidence on social investing from USAID’s development innovation ventures. 2019;

23. Kniesner TJ, Viscusi WK. The value of a statistical life. Oxford Research Enyclopedia of Economics and Finance. Vanderbilt Law Research Paper No. 19-15; 2019.

24. Zaigham M, Varallo J, Thangaratinam S, Nicholson W, H. A. Visser G (2024) Global disparities in caesarean section rates: Why indication-based metrics are needed. PLOS Glob Public Health 4(2): e0002877

25. MI, Ryan GF, Britt DW, Macedonia CR: The Mortality of Politics: An American paradox. Fetal Diagn Therapy 2024 10.1159/000541912

26. Evans MI, Britt DW, Devoe LD. Etiology and ontogeny of cerebral palsy: implications for practice and research. Reprod Sci. 2024;31(5):1179–1189. doi:10.1007/s43032-023-01422-6 20a.

27. Evans MI: Overcoming Militant Mediocrity Am J Obstet Gynecol 2008; 198:656–661

28. Evans MI, Britt DW, Evans SM, Devoe LD: Improving the interpretation of electronic fetal monitoring: the fetal reserve index. Am J Obstet Gynecol 2023;228:S1129–1143

29. Levitt L, Altman D. Complexity in the US health care system is the enemy of access and affordability. JAMA Health Forum. 2023;4(10):e234430–e234430. doi:10.1001/jamahealthforum.2023.4430

30 Philbbs CM, Kristensen-Cabrera A, Kozhimannil KB, Leonard SA, Lorch SA, Main EK, et al: Racial/ethnic disparities in costs, length of stay,and severity of severe maternal morbidity. Am J Obstet Gynecol MFM 2023;5:100971 doi: 10.1016/j.ajogmf.2023.100917O

31 Mujahid MS, Wall-Wieler E, Hailu EM, Berkowitz RL, Gao X, Morris CM, et al: Neighborhood disinvestment and severe maternal maternal morbidity in thestate of California. Am J Obstet Gynecol MFM 2023; doi: 10.1016/j.ajogmf.2023.100916

32. Cole MB, Kim J, Gordon SH, Lasser KE, Ncube C, Patton E, et al: Massachusetts Medicaid ACO Program May Have Improved Care Use And Quality For Pregnant And Postpartum Enrollees, Health Aff 2024;43.00230

33. Neumann PJ, Kim DD. Cost-effectiveness thresholds used by study authors, 1990-2021. JAMA. 2023;329(15):1312–1314. doi:10.1001/jama.2023.1792

34 Cerf ME: The social-education-economy-health nexus, development and sustainability: perspectives from low and middle-income and African countires. Discov Sustain 2023; 10.1007/s43621-023-00153-7

35. Caraballo C, Massey DS, Ndumele CD, et al. Excess mortality and years of potential life lost among the black population in the US, 1999-2020. JAMA. 2023;329(19):1662–1670. doi:10.1001/jama.2023.7022

36 Evans MI, Britt DW, Evans SM: Midforceps did not cause “compromised babies” – compromise caused forceps: an approach toward safely lowering the cesarean delivery rate. J Matern Fetal Neonat Med 2022; 29:1874–1894

37. Jiang HJ, Moshe Henke R, Fingar KR, Denis Agniel LL, Roemer MI: Rural hospitals experienced more patient volume variability than urban hospitals during the COVID-19 Pandemic, 2020-21. Health Aff 2024;43 (5)

38. Henke RM, Fingar KR, Jiang HJ, Liang L, Gibson TB. Access to obstetric, behavioral health, and surgical inpatient services after hospital mergers in rural areas. Health Aff (Millwood). 2021;40(10):1627– 36.

39. Polan RM, Barber EL. Reimbursement for female-specific compared with male-specific procedures over time. Obstet Gynecol. 2021;138(6):878–883. doi:10.1097/AOG.0000000000004599

40. https://www.healthsystemtracker.org/chart-collection/health-expenditures-vary-acrosspopulation/#Share%20of%20total%20population%20and%20total%20health%20spending,%20by%20age%20group,%202021)

41. U.S. Social Security Administration. Monthly Statistical Snapshot: January 2024. https://www.ssa.gov/policy/docs/quickfacts/stat_snapshot/index.html

42. Commonwealth of Massachusetts. Special Education Tuition Pricing Details. 26. https://www.mass.gov/info-details/special-education-tuition-pricing-details

43. Peters T: Re-Imagine! Business Excellence in a Disruptive Age. DK Publishing, NY 2003.

44. Evans MI, Britt DW: Resistance to Change Reprod Sci: 2023;30:835–853

45. Batman S, Rivlin K, Robinson W, Brown O, Carter EB, Lindo E: A rubric to center equity in obstetrics and gynecology research. Obstet Gynecol 2023;142:772–778

46. Doll KM: The future of research on racism in reproductive health. Obstet Gynecol 2023;142:987–988.

47. Henke RM, Fingar KR, Jiang HJ, Liang L, Gibson TB. Access to obstetric, behavioral health, and surgical inpatient services after hospital mergers in rural areas. Health Affairs. 2021;40(10):1627–1636. doi:10.1377/hlthaff.2021.00160

48. Pineda R, Kati K, Breault CC, Rogers EE, Mack WJ, Fernandez-Fernandez A. NICUs in the US: levels of acuity, number of beds, and relationships to population factors. J Perinatol. 2023/06/01 2023;43(6):796–805. doi:10.1038/s41372-023-01693-6

49. https://www.bcbs.com/the-health-of-america/articles/healthy-communities-mean-better-economy

50. Policy Framework March 2023. US Agency for international development. https://www.usaid.gov/policy/documents/mar 23-2023.

51. Chappel, A., DeLew, N., Grigorescu, V., & Smith, S. R. (2021). Addressing the maternal health crisis through improved data infrastructure: guiding principles for progress. Health Affairs Forefront.

52. Peahl, A. F., & Shah, N. (2023). The Maternal Health Crisis Is A Consequence Of Design. Health Affairs Forefront.

53. Attanasio, L. B., & Geissler, K. H. (2024). The Role Of Medicaid Accountable Care Organizations In Maternal Health. Health Affairs Forefront.

54. Bowser DM, Maurico K, Ruscitti BA, Crown WH: American clusters: using machine learning to understand health and healthcare disparities in the United States. Health Aff Scholar 2024;2 qxae017.

